# Assessment of COVID-19 vaccine acceptance among healthcare workers in Los Angeles

**DOI:** 10.1101/2020.11.18.20234468

**Authors:** Adva Gadoth, Megan Halbrook, Rachel Martin-Blais, Ashley Gray, Nicole H. Tobin, Kathie G. Ferbas, Grace M. Aldrovandi, Anne W. Rimoin

## Abstract

**Importance:** Healthcare workers (HCW) are slated to be early recipients of SARS-CoV-2 vaccines due to increased risk of exposure to patients with COVID-19, and will be tasked with administering approved vaccines to the general population. As lynchpins of the vaccination effort, HCWs’ opinions of a vaccine’s safety and efficacy may affect both public perception and uptake of the vaccine. Therefore, it is crucial to understand and address potential hesitancy prior to vaccine administration.

**Objective:** To understand healthcare workers’ attitudes about vaccine safety, efficacy, and acceptability in the context of the COVID-19 pandemic, including acceptance of a novel coronavirus vaccine.

**Design, Setting, Participants:** A cross-sectional survey was distributed to participants enrolled in a longitudinal cohort study surveilling SARS-CoV-2 infection among 1,093 volunteer sampled University of California, Los Angeles (UCLA) Health System employees. Surveys were completed online between September 24 and October 16, 2020. In total, 609 participants completed this supplemental survey.

**Results:** We averaged a 9-statement Likert scale matrix scored from 1 (“strongly disagree”) to 5 (“strongly agree”) and found respondents overwhelmingly confident about vaccine safety (4.47); effectiveness (4.44); importance, self-protection, and community health (4.67). Notably, 47.3% of respondents reported unwillingness to participate in a coronavirus vaccine trial, and most (66.5%) intend to delay vaccination. The odds of reporting intent to delay coronavirus vaccine uptake were 4.15 times higher among nurses, 2.45 times higher among other personnel with patient contact roles, and 2.15 times higher among those without patient contact compared to doctors. Evolving SARS-CoV-2 science (76.0%), current political climate (57.6%), and fast-tracked vaccine development timeline (83.4%) were cited as primary variables impacting HCW decisions to undergo vaccination. Of note, these results were obtained prior to release of Phase III data from companies manufacturing vaccines in the U.S.

**Conclusions and Relevance:** Despite overall confidence in vaccines, a majority of HCW expressed concerns over a novel coronavirus vaccine. A large proportion plan to delay vaccine uptake due to concerns about expedited development, emerging scientific discoveries, and the political climate. Forthcoming vaccination campaigns must address these unique points of coronavirus vaccine hesitancy in order to achieve adequate vaccine coverage.

## Background

Vaccines prevent disease, disability, and death for millions annually, yet public support for their use has been waning worldwide. The rise of vaccine hesitancy, including the delay or refusal of immunization, poses real and existential threats to progress against vaccine-preventable disease outbreaks, and is increasingly being recognized as a barrier to immunization program success.^1,2^ Indeed, in 2019 the World Health Organization listed vaccine hesitancy as one of the top ten threats to global health.^3^

Long-term coronavirus disease (COVID-19) control will likely hinge on successful vaccine development and delivery to a large portion of the population in order to achieve adequate vaccine coverage to prevent ongoing community spread. The highly politicized U.S. governmental pandemic response has ignited concerns that public acceptance of a novel coronavirus vaccine may be insufficient to establish herd immunity as a result of mistrust of authorities, misinformation on the internet, and other sources of vaccine hesitancy. Indeed, several recent surveys in the U.S. and abroad have shown mass uncertainty regarding vaccines and the influence of modern political movements,^4^ with implications for coronavirus vaccine acceptance.^5,6^ Skepticism over the fast-tracked vaccine development and approval process known as Operation Warp Speed, also has the public worried that politics rather than science might be driving a vaccine to market.^7-9^

Studies in high-, middle-, and low-income settings have consistently shown that healthcare workers (HCW) offer guidance on vaccine recommendations and help combat misinformation, confusion, and ignorance about the risks and benefits of vaccination to the public.^10^ Nonetheless, blanket HCW support for vaccines should not be taken for granted. A recent study found that 43% of health practitioners in France did not recommend vaccines to key demographic groups, and carried strong perceptions of vaccine risk based on decades-old societal controversies.^11^ Similar beliefs color health practitioner vaccine behaviors around the world, and underscore the fact that vaccine hesitancy is often influenced by broader political, religious, social, and historical factors.^12^

To understand the extent of generalized vaccine acceptance and specific attitudes towards forthcoming coronavirus vaccines among HCW in Los Angeles, we conducted a cross-sectional survey delivered to health system employees enrolled in a longitudinal SARS-CoV-2 surveillance program.

## Methods

### Study design and population

To understand the risk of SARS-CoV-2 infection among healthcare workers^13^ we enrolled a cohort of asymptomatic healthcare workers employed by University of California, Los Angeles (UCLA) Health at two medical centers into a longitudinal study from April 8 to October 14, 2020. Participants were asked to complete biweekly mid-turbinate swab sampling for SARS-CoV-2 testing via nucleic acid amplification, and to provide blood samples once per month to assess the presence of anti-SARS-CoV-2 IgG. Subjects also completed an online baseline questionnaire on basic demographics, as well as a biweekly survey on recent occupational exposures, including contact with patients or biologic samples that had tested positive for SARS-CoV-2.

### Survey design

As an addendum to this study, a brief cross-sectional survey designed to assess attitudes towards vaccines and prospective acceptance of a novel coronavirus vaccine was distributed to participants on September 24, 2020 and completed online through October 16, 2020. Survey questions were modeled on validated vaccine questionnaires published as part of the World Health Organization (WHO) Strategic Advisory Group of Experts (SAGE) on Immunization’s vaccine hesitancy Likert scale questions^14,15^, and a survey on public perceptions of COVID-19 from the Vaccine Confidence Project.^16^

Nine Likert scale questions from SAGE were modified to examine attitudes about vaccines at large, rather than childhood vaccination (Table 1). Answers to each statement were assigned a point value from 1-5 (from “strongly disagree” to “strongly agree”). Additional questions were incorporated to address prospective acceptance of a novel coronavirus vaccine; factors influencing participants’ opinions of such a vaccine, including political, social, and religious variables; and which groups, if any, they believe should have priority access. Subjects were also asked to reflect on how the COVID-19 pandemic has impacted their willingness to vaccinate.

**Table 1.** Average Likert Response by Demographic Variable

| Survey Questions | Education |  |  |  |  |
| --- | --- | --- | --- | --- | --- |
| | Some College, Associate's, or Vocational Degree | Bachelor's Degree | Advanced Degree | p-value | $\beta$ coefficient and 95% CI |
| 1 Vaccines are important for my health. | 4.30 | 4.52 | 4.81 | <.0001 | 0.266 (0.198, 0.336) |
| 2 Getting vaccines is a good way to protect myself from disease. | 4.27 | 4.54 | 4.80 | <.0001 | 0.257 (0.19, 0.324) |
| 3 Overall, vaccines are safe. | 4.06 | 4.26 | 4.67 | <.0001 | 0.361 (0.282, 0.441) |
| 4 Overall, vaccines are effective. | 4.04 | 4.22 | 4.62 | <.0001 | 0.329 (0.252, 0.406) |
| 5 Getting vaccinated is important for the health of others in my community. | 4.24 | 4.56 | 4.81 | <.0001 | 0.273 (0.211, 0.336) |
| 6 The information I receive about vaccines from public health authorities/ my healthcare provider is reliable and trustworthy. | 3.90 | 4.07 | 4.36 | <.0001 | 0.250 (0.154, 0.346) |
| 7 Generally, I do what my doctor or health care provider recommends about vaccines for myself and my family. | 4.02 | 4.26 | 4.59 | <.0001 | 0.309 (0.225, 0.393) |
| 8 New vaccines carry more risk than older vaccines. | 3.30 | 3.39 | 3.15 | 0.04 | -0.131 (-0.002, -0.26) |
| 9 I am concerned about serious adverse effects of vaccines. | 3.52 | 3.18 | 2.71 | <.0001 | -0.432 (-0.295, -0.57) |

p-values obtained from two-sample t-tests and ANOVAs.
Linear trend assessed via crude linear regression.
Likert values used for averages: 1- Strongly Disagree, 2- Disagree, 3- Neutral, 4- Agree, 5- Strongly Agree

### Statistical analysis

Descriptive statistics were calculated for demographic factors of both the greater study cohort and those who opted to complete the vaccine hesitancy survey (Table 2). Answers to the vaccine hesitancy Likert scale questions were tabulated, and then averaged for the whole group and for specific demographic categories. T-tests or ANOVAs were also run to detect meaningful differences in answers across these groups, with linear regression conducted to examine trends across ordinal education levels.

**Table 2.** Participant Demographics

|  | **Vaccine Hesitancy (N=609)** |  | **All (N= 1093)** |  |
| --- | --- | --- | --- | --- |
|  | **n** | **col %** | **n** | **col%** |
| **Age** |  |  |  |  |
| 18-29 | 89 | 14.61 | 178 | 16.29 |
| 30-39 | 230 | 37.77 | 438 | 40.07 |
| 40-49 | 136 | 22.33 | 245 | 22.42 |
| 50-59 | 88 | 14.45 | 149 | 13.63 |
| 60 + | 42 | 6.9 | 59 | 5.40 |
| Missing | 24 | 3.94 | 24 | 2.20 |
| **Sex** |  |  |  |  |
| Male | 163 | 26.77 | 327 | 29.92 |
| Female | 419 | 68.8 | 735 | 67.25 |
| Missing | 27 | 4.43 | 31 | 2.84 |
| **Race\*** |  |  |  |  |
| Asian | 166 | 27.26 | 318 | 28.49 |
| American Indian/Alaska Native | 7 | 1.15 | 9 | 0.81 |
| Black/African American | 23 | 3.78 | 47 | 4.21 |
| Native Hawaiian/other Pacific Islander | 13 | 2.13 | 34 | 3.05 |
| White | 345 | 56.65 | 586 | 52.51 |
| Other | 47 | 7.72 | 98 | 8.78 |
| Missing | 24 | 3.94 | 24 | 2.15 |
| **Ethnicity** |  |  |  |  |
| Hispanic/Latino | 69 | 11.33 | 143 | 13.08 |
| Non-Hispanic/Latino | 472 | 77.5 | 818 | 74.84 |
| Prefer not to say | 25 | 4.11 | 55 | 5.03 |
| Missing | 43 | 7.06 | 77 | 7.04 |
| **Education** |  |  |  |  |
| Graduated high school/obtained GED | 3 | 0.49 | 9 | 0.82 |
| Some college | 50 | 8.21 | 102 | 9.33 |
| Bachelor's degree | 168 | 27.59 | 333 | 30.47 |
| Advanced degree | 363 | 59.61 | 620 | 56.72 |
| Missing | 25 | 4.11 | 29 | 2.65 |
| **Household Income** |  |  |  |  |
| Less than or equal to \$25,000 | 6 | 0.99 | 12 | 1.10 |
| \$25,000 to \$50,000 | 12 | 1.97 | 25 | 2.29 |
| \$50,001 to \$75,00 | 74 | 12.15 | 134 | 12.26 |
| \$75,001 to \$100,000 | 66 | 10.84 | 145 | 13.27 |
| \$100,001 to \$125,000 | 81 | 13.3 | 152 | 13.91 |
| \$125,001 to \$150,000 | 74 | 12.15 | 118 | 10.80 |
| Over \$150,000 | 240 | 39.41 | 420 | 38.43 |
| Don't know | 2 | 0.33 | 4 | 0.37 |
| Prefer not to say | 30 | 4.93 | 56 | 5.12 |
| Missing | 24 | 3.94 | 27 | 2.47 |

\* Participants were allowed to select more than one race

Attitudes surrounding a novel, as-of-yet unspecified, coronavirus vaccine were graphed. Proportions of subjects intending to accept, delay, or refuse a future vaccine were calculated, as were the motivating factors behind these decisions. All vaccine attitudes were stratified by sex, race, ethnicity, and level of educational attainment. Finally, a logistic regression model was run to examine the relationship between prospective COVID-19 vaccine receipt and various demographic factors of interest. All analyses were performed using SAS version 9.4 (Cary, NC) and R (R Core Team, 2014) statistical software; figures were produced using the R package ggplot2 (Wickham, 2009).

### Ethical considerations

Ethical approval for this study was obtained from the UCLA Institutional Review Board (IRB #20-000478).

## Results

### Study population

Among 1,069 participants in our healthcare worker cohort, 609 (55.7%) opted to complete a vaccine hesitancy questionnaire. Similar to the larger cohort, these participants were majority female (68.8%), white (56.7%), between the ages of 30-49 (60.1%) and in possession of an advanced degree (59.6%, Table 2). Vaccine module respondents also mirrored the larger HCW cohort in terms of job role, with 39.9% physicians, nurse practitioners, physician assistants, or certified registered nurse anesthetists (CRNAs); 33.8% nurses; 16.6% in other patient contact roles in the health system; and 9.8% personnel without patient contact.

### General vaccine attitudes

When asked about attitudes towards nine statements designed to measure acceptance of vaccines at large, respondents were overwhelmingly positive. Ninety-three percent (range: 84.0% - 96.9%) surveyed agreed or strongly agreed with Statements 1-7 (listed in Table 1), indicating high vaccine confidence (average response of 4.51/5 on the Likert scale). 85.7% (n=522) of participants responded ‘agree’ or ‘strongly agree’ to all seven statements; 33.5% (n=204) responded ‘strongly agree’ to all seven statements.

Responses to statements designed to assess vaccine hesitancy (Statements 8-9, also listed in Table 1) were less positive than those that measured acceptance of vaccines at large (Statements 1-7). The average response to the statement ‘new vaccines carry more risk than older vaccines’ was 3.23/5 and the average response to ‘I am concerned about serious adverse effects of vaccines’ was 2.9/5. Over one third of participants (35.0%) expressed concern about serious adverse effects from vaccine use; almost a quarter (23.3%) were neutral on the issue.

The average Likert response was higher among men compared with women for statements regarding the safety and effectiveness of vaccines (Statements 2-4) and the importance of vaccines for community health (Statement 5). A greater proportion of men strongly agreed with Statements 1-7 compared with women, and participants identifying as Latino were less likely to respond ‘strongly agree’ compared with non-Latinos. When asked whether they trusted vaccination information provided by healthcare and public health professionals, 17.4% of Latino respondents were neutral, 50.7% agreed that this information was trustworthy, and 29% strongly agreed, compared with 11.9%, 39.2% and 45.8% of non-Latinos, respectively.

Nearly all of our participants (95.4%) reported either some college, associate’s, or a vocational degree; completion of a bachelor’s degree; or attainment of an advanced degree. We observed a linear, dose-response trend in vaccine acceptance and hesitancy across these three levels of education. We observed a positive linear trend signaling increasing vaccine confidence (Statements 1-7) with increased educational attainment, and a negative linear trend signaling reduced vaccine hesitancy (Statements 8-9) with increased education level.

### Attitudes towards novel coronavirus vaccine

Participants were asked about a prospective novel coronavirus vaccine during a period when many vaccine candidates were under development globally, and several had advanced to Phase III testing in the United States. Almost half of participants (46.0%) agreed that a coronavirus vaccine would protect them from COVID-19 disease, 48.4% responded neutrally, and 5.60% disagreed. Among Latinos, 27.5% agreed that a novel coronavirus vaccine would protect them from COVID-19, 65.2% responded neutrally, and 7.25% disagreed. Confidence in the scientific vetting process of a novel coronavirus vaccine dropped markedly and did not differ according to ethnicity, with 37.6% of participants expressing neutrality on the matter, and 27.8% stating they were not confident in these procedures. Finally, almost half of respondents (47.3%) stated they were unwilling to participate in a novel coronavirus vaccine trial (Figure 1).

**Figure 1.**
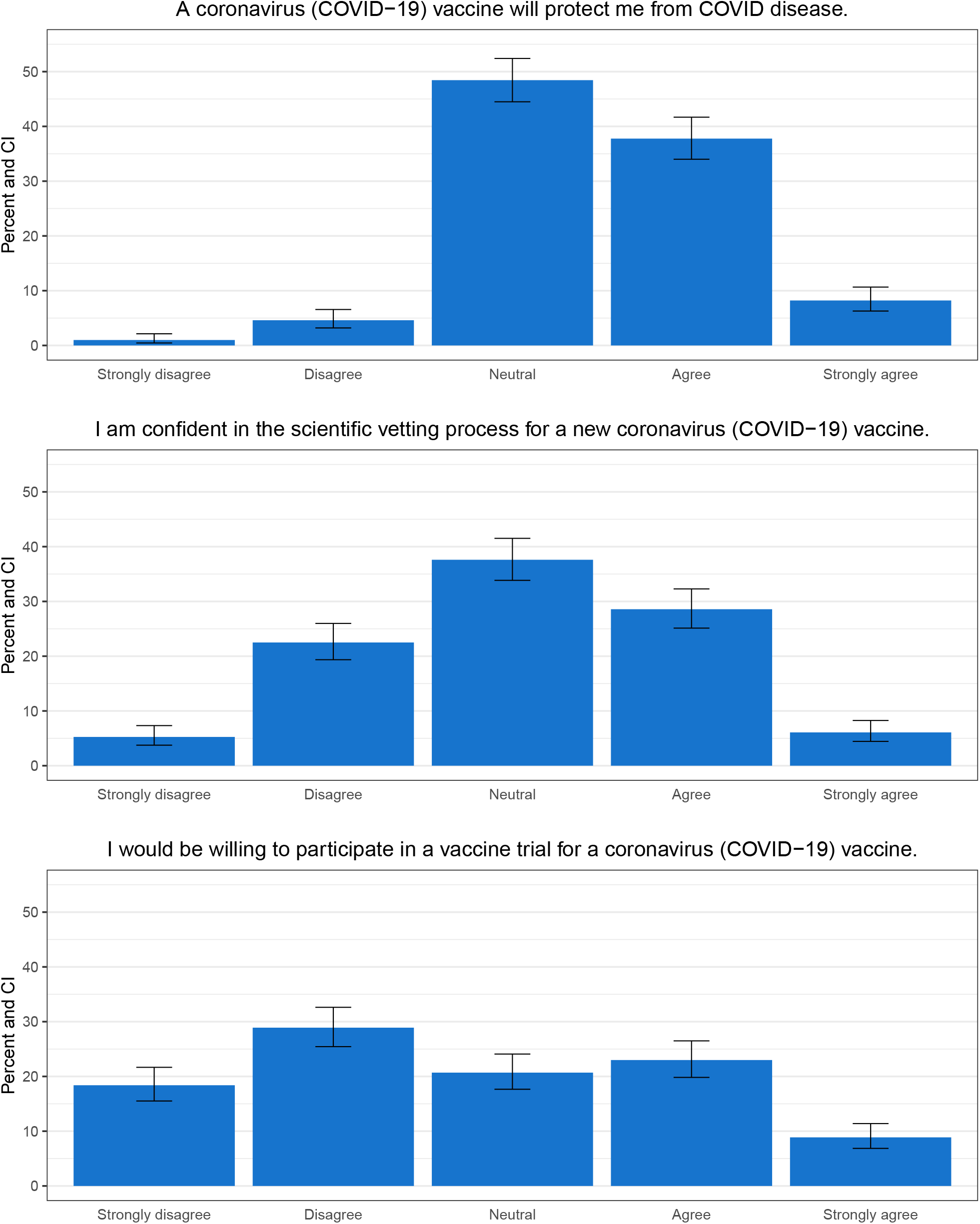
Participant attitudes regarding a coronavirus vaccine

Of importance, most participants (66.5%) indicated that they would delay vaccination if a new coronavirus vaccine became widely available: 49.9% would prefer to wait and see how the vaccine affects others first and 16.6% would not get it soon, but indicated they might in the future; 1.31% (n=8) never intend to get vaccinated. The remaining participants (32.3%) reported that they intended to get a coronavirus vaccine as soon as possible. We fit a logistic regression model to measure the association between coronavirus vaccine intent and demographic factors of interest (Table 3). Compared with physicians, nurse practitioners, physician assistants and CRNAs, other HCW had significantly lower odds of coronavirus vaccine acceptance. The odds of delay or refusal of a coronavirus vaccine was 4.17 times higher (95% CI: 2.53-6.87) among nurses, 2.71 times higher (95% CI: 1.51-4.84) among other personnel with patient contact roles, and 1.71 times higher (0.87-3.34) among those with other roles in the health system that did not include patient contact.

**Table 3.** Coronavirus intent-odds of delaying coronavirus vaccine

|  | Variable & Wald p-value | aOR | 95% CI |
| --- | --- | --- | --- |
| Job Role<br><.0001 | Doctors<sup>1</sup> | *ref* | *ref* |
|  | Nurses | 4.17 | (2.53, 6.87) |
|  | Other personnel with direct patient contact | 2.71 | (1.51, 4.84) |
|  | Personnel without patient contact | 1.71 | (0.87, 3.34) |
| Sex<br>0.2643 | Male | *ref* | *ref* |
|  | Female | 1.28 | (0.83, 1.98) |
| Race<br>0.0245 | White | *ref* | *ref* |
|  | Asian | 2.28 | (1.40, 3.71) |
|  | Black | 1.47 | (0.42, 5.15) |
|  | Other | 1.22 | (0.56, 2.66) |
|  | Two or more | 1.06 | (0.45, 2.49) |
| Ethnicity<br>0.1609 | non-Latino | *ref* | *ref* |
|  | Latino | 1.66 | (0.82, 3.39) |

aOR= adjusted odds ratio
<sup>1</sup> Includes doctors, nurse practitioners, physician assistants, and CRNAs

Those planning to delay or refuse vaccination cited concerns regarding the fast-tracked nature of vaccine development and a lack of transparency and/or publicly available information on a vaccine as their primary rationale. Those factors were noted again amongst the survey respondents at large, regardless of vaccination timing preferences. Opinions guiding intentions for a potential coronavirus vaccine were most heavily influenced by the fast-tracked development timeline, the novel and unfolding science of SARS-CoV-2, and the political climate in which the research and regulatory process were playing out at the time of survey distribution (Figure 2). These three concerns persisted across different demographic groups including, sex, ethnicity, and education level.

**Figure 2.**
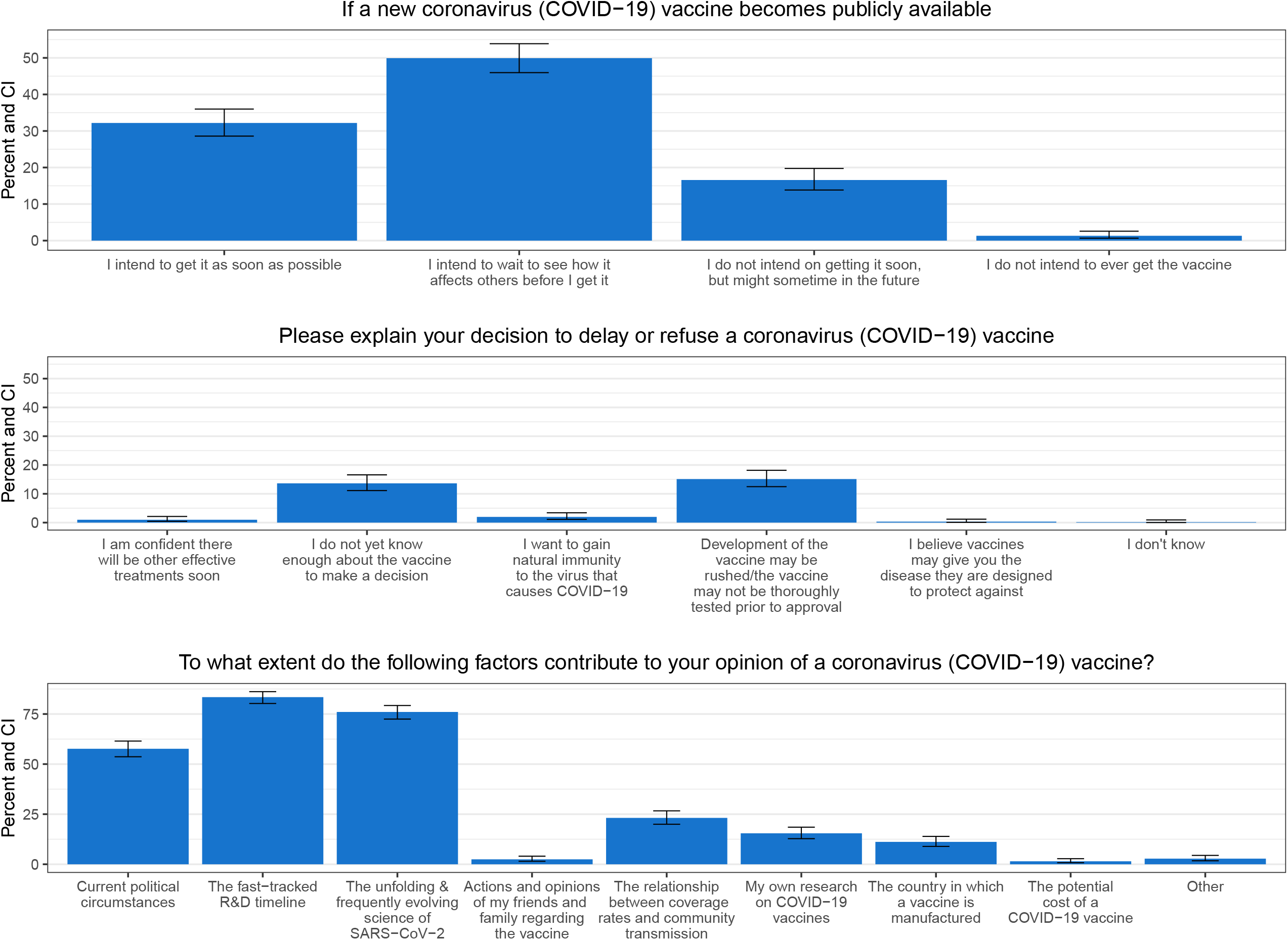
Participant intentions regarding a coronavirus vaccine

When asked if the COVID-19 pandemic had impacted their willingness to be immunized in general, 70.1% of subjects reported no change, 20.9% felt somewhat more likely or much more likely to receive a vaccine, and 9.04% felt somewhat or much less likely to receive a vaccine. After being shown a comparison of the reproductive numbers for SARS-CoV-2 (R_0_∼ 3) and measles (R_0_∼ 14), participants were asked about their willingness to accept measles-containing vaccines. Over three-quarters (77.2%) said the relative infectiousness of measles disease led to no change in their feelings around vaccination, 20.0% felt somewhat more likely or a lot more likely to accept measles-containing vaccines, and 2.8% felt somewhat or a lot less likely to vaccinate against measles.

## Discussion

As arbiters of scientific and public health information, healthcare workers serve as ambassadors for evidence-based medical interventions, and are critical in promoting vaccine acceptance.^17^ That has never been more important than in the wake of COVID-19. Not only do HCW serve on the frontlines of pandemic response efforts, at high risk for occupational SARS-CoV-2 exposure and transmission, but they are almost universally identified as priority recipients of a forthcoming coronavirus vaccine.^18^ HCW also serve as trusted and influential community members on topics of public health concern, and will ultimately be prescribing and administering approved coronavirus vaccines to their patients. While our study population generally regarded vaccines very positively and indicated high willingness to follow public health guidelines around vaccination, they expressed some skepticism and specific concern over a COVID-19 vaccine.

Although participants overwhelmingly acknowledged the safety, efficacy, and importance of vaccination to public health practice, 35% expressed apprehension over serious adverse effects from vaccines, and 23% said they were “neutral” with respect to serious adverse effects. While it’s impossible to precisely define “neutral” responses in this context, this could indicate a lack of assuredness over the perceived safety of all vaccines, or may suggest that respondents make vaccination decisions on a vaccine-by-vaccine basis.

Notably, we observed several instances where women and racial or ethnic minorities expressed more hesitant attitudes towards vaccines than their counterparts. Persistent issues of trust in the modern medical system among these groups speaks to a larger trend in perceptions of healthcare among minority groups in the U.S., which may drive individuals to seek alternative or complimentary medicine, or to neglect health concerns altogether.^19-21^ This trust gap, which stems in part from long-term issues of systemic biases in health research and healthcare delivery^22,23^, will have important implications for the adoption and roll-out of a novel coronavirus vaccine.

At the time of survey distribution in late September of 2020, hesitancy over an upcoming coronavirus vaccine in our cohort appeared to be grounded in a lack of confidence about the timeline and motives pushing such vaccines to market. Several polls and surveys of the adult U.S. population from this same time period show concern surrounding ongoing Phase III trials of several vaccine candidates, owing to perceived lack of data clarity, financial and political conflicts of interest, and other current events playing out in the news cycle.^7-9^ A perceived lack of transparency in government-supported vaccine development programs and regulatory decision-making has further perpetuated and broadened public mistrust, including messaging around the public-private partnership known as Operation Warp Speed, which to some connotes “cutting corners” to rapidly advance vaccines to market.^24^ As a reflection of these collective public sentiments, an alliance of governors from Western states and New York state are vowing to conduct secondary scientific reviews of any federally approved vaccines before distributing them to their residents.^25^

In the context of healthcare uptake and delivery, apparent meddling of the government in the scientific process of vaccine development may hamper enthusiasm around novel interventions without a long history of post-market surveillance supporting their use. This may help explain why two-thirds of respondents wanted to delay use of a novel coronavirus vaccine, some with no intention of getting the vaccine in the immediate future. Alternatively, the perceived risk of COVID-19 may be lower for young, generally healthy individuals in the workforce than the perceived risk of adverse effects from a coronavirus vaccine. For individuals under age 50 without comorbidity, the risk of death from COVID-19 remains low, and up to 40% have completely asymptomatic disease.^26^ HCWs may thus be more hesitant to receive a novel vaccine when they do not perceive the disease it protects against as a significant personal threat.

Hesitancy around uptake of a coronavirus vaccine was greater among nurses than among physicians and physician counterparts. This trend has been shown in numerous studies of attitudes around influenza vaccination, and should be taken seriously by public health authorities considering the fact that nurses frequently interface with patients, particularly around vaccine administration, and often are in charge of directly administering vaccines.^27-29^

In the context of forthcoming distribution efforts, in which healthcare workers have frequently and publicly been touted as priority vaccine recipients^18^, these findings seem to suggest that many in the field might be wary of serving as “guinea pigs” for vaccines that have had limited public messaging around their effectiveness, side effects, and other parameters of interest to this group. Despite this, 69.3% of our cohort still felt that healthcare workers should have early access to a vaccine. These apparent contradictions may be explained by a desire to ensure vaccine accessibility while maintaining decisional autonomy over one’s personal healthcare. They may also reflect the inherent role that emotions, personal values, and worldviews play in health decision making,^30-32^ a key facet for public health communicators to take into account when marketing novel vaccines to both the healthcare community and the general public.^33^

This study had several limitations. Although the majority of participants opted in to this supplemental vaccine attitudes survey, a high proportion of cohort members did not. Despite this, a side-by-side comparison of the full study cohort and vaccine module respondents shows similar demographic breakdowns for each, diminishing the risk of selection bias. Our cohort was overwhelmingly highly educated and of higher socioeconomic status. This trend is, in part, a reflection of the post-graduate degrees required by those in many healthcare positions, though a more thorough sampling of other health system employees with diverse economic backgrounds and job types should be pursued for recruitment in future studies. Additionally, this cross-sectional study measured vaccine attitudes at a specific time point in the COVID-19 pandemic from late September to mid-October, 2020. As major vaccine news, political circumstances, and regional epidemiologic data change on a daily basis, it is likely that vaccine attitudes will change frequently over time, and thus will need to be longitudinally monitored.

As national and local experiences during the COVID-19 pandemic have unveiled our deficiencies in public health communication, HCW remain key allies in disease control strategies. Hesitancy towards a novel coronavirus vaccine among this cohort should serve as a warning sign to public health authorities, as it could trigger a ripple effect in the general public. It may also signal an unwillingness on the part of health practitioners to administer a vaccine to patients in later stages of vaccine distribution, once more doses are available.^10,34^

A public health-led coronavirus vaccine roll-out will need to address concerns raised by the public and HCW regarding the ‘warp speed’ research and development timeline, lack of publicly available data from ongoing vaccine trials, and the highly politicized vaccine approval process if it hopes to gain broad acceptance and public buy-in. Our survey found that being confronted with simple scientific facts regarding disease transmission, like a reproductive number, can increase one’s willingness to vaccinate. Future inquiries on this topic should examine the politicization of vaccine development and whether results of the 2020 U.S. Presidential election have in any way modified the likelihood of coronavirus vaccine acceptance. Examining the impact of science-based marketing strategies will also be key to this effort. A recent study on coronavirus vaccine attitudes suggests reporting higher rates of vaccine effectiveness is correlated with greater public vaccine acceptance.^35^ Publicized, data-driven updates from ongoing vaccine trials may have a positive effect on healthcare worker willingness to receive and recommend coronavirus vaccines. Successful rollout of a novel coronavirus vaccine will need to build trust and confidence in communities across the country and leverage public health allies—and it must start with healthcare workers.

## Data Availability

Requests for data should be submitted to the corresponding author.

## Acknowledgements

The authors would like to thank Dr. Heidi J. Larson and the Vaccine Confidence Project for sharing survey questions on public perceptions of a COVID-19 vaccine that were incorporated into our questionnaire. We would also like to thank the UCLA Health employees who participated in our study.

